# Emergence and rising of ceftazidime-avibactam resistant KPC-producing *Pseudomonas aeruginosa* in China: a molecular epidemiology study

**DOI:** 10.1101/2020.12.15.20248268

**Authors:** Yiwei Zhu, Jie Chen, Han Shen, Zhongju Chen, Qi-wen Yang, Jin Zhu, Xi Li, Qing Yang, Feng Zhao, Jingshu Ji, Heng Cai, Yue Li, Linghong Zhang, Sebastian Leptihn, Xiaoting Hua, Yunsong Yu

## Abstract

**Background:** Infections by Carbapenem-resistant *Pseudomonas aeruginosa* (CRPA) are difficult to treat and novel antibiotics are desperately needed. Till today, ceftazidime-avibactam (CAZ-AVI) has been used to treat infections caused by multidrug resistant (MDR) Gram-negative bacteria, including *Klebsiella pneumoniae* carbapenemase (KPC)-producing organisms. Although cases of KPC-producing *P. aeruginosa* (KPC-PA) have been reported sporadically, data about KPC-PA susceptibility to CAZ-AVI and its molecular characteristics are limited.

**Methods:** CRPA were collected from seven hospitals in China from 2017 to 2018. PCR was deployed to screen for the *bla*_KPC_ gene. Antimicrobial susceptibility of KPC-PA was determined by broth microdilution method or agar dilution method. We combined Illumina sequencing and Nanopore long-read sequencing to elucidate the genomic characteristics of KPC-PA strains.

**Results:** KPC-PA strains were found in six out of seven hospitals. 151/374 (40.4%) CRPA clinical isolates harbored the *bla*_KPC-2_ gene. Among KPC-PA, ST463 (107/151) was predominant, followed by ST485 (14/151) and ST1212 (12/151). Approximately half of all KPC-PA (50.3%) were susceptible to CAZ-AVI. We found that the *bla*_KPC-2_ gene copy number correlated with CAZ-AVI MIC. In more than 90% (136/151) of the strains, we found plasmids that belonged to two types carrying *bla*_KPC-2_ gene. The Type 1 plasmid, predominant in East China, was identified in 118 strains and the Type 2 plasmid, belonged to a megaplasmid family spreading globally, was identified in 19 strains. In addition, we identified IS*26*-ΔTn*6296* and IS*6100*-ΔTn*6296*-Tn*1403* as two mobile genetic elements that mediated *bla*_KPC-2_ gene transmission.

**Conclusion:** Our results suggest that the *bla*_KPC-2_ gene is becoming a remarkable mediator of carbapenem resistance in *P. aeruginosa* in China. The emergence and spread of such KPC-PA strains poses a threat on clinical therapy as CAZ-AVI becomes inefficient. It would be beneficial to screen for the *bla*_KPC_ gene in CRPA strains for antimicrobial surveillance.

## Background

*Pseudomonas aeruginosa*, one of major opportunistic pathogens, is notorious for its potency to resist antibiotics including carbapenems. Although carbapenems are still among the first-line therapeutics for infections caused by multi-drug resistant (MDR) *P. aeruginosa*, carbapenem-resistant *P. aeruginosa* (CRPA) are increasingly observed in the clinic. Indeed, CRPA is among the pathogens listed by the World Health Organization (WHO) that are considered of high relevance for human health and for which new antibiotics or clinical strategies are urgently needed [1].

Approved by the U.S. Food and Drug Administration (FDA) in 2015, ceftazidime-avibactam (CAZ-AVI), a novel β-lactam/β-lactamase inhibitor combination, has been deployed in the clinic for complicated intraabdominal infections and for hospital-acquired pneumonia caused by multidrug-resistant *Enterobacteriaceae* and *P. aeruginosa* [2]. The inhibition spectrum of avibactam includes the *Klebsiella pneumoniae* carbapenemase family (KPC) [3]. According to INFORM surveillance [4-6], CAZ-AVI susceptibility rates are between 84-90% to CRPA that do not express metallo-β-lactamase (MBL). However, no study has detailed the CAZ-AVI susceptibility of KPC-PA as a single group, and investigated occurring resistance mechanisms, possibly due to the currently low prevalence of such strains. Perhaps unsurprisingly, an increasing number of *bla*_KPC_ genes have been detected in clinical *P. aeruginosa* isolates over the last years [7-12]. An example is the Eastern Chinese city of Hangzhou, where the rate of KPC-PA has increased significantly since the first case was reported a decade ago [13, 14].

Most of the investigated *bla*_KPC_ genes in *P. aeruginosa* have been reported to be plasmid-encoded [15-18], while a small number are found on the bacterial chromosome, occasionally in prophage sequences [12, 19]. However, most of these studies are single case reports and focus on the description of the genetic structure of *bla*_KPC_ and its surrounding sequences. Therefore, the importance of KPC-PA strains and their relevance for the clinic remain unclear.

In this study, we analyzed the prevalence of KPC-PA strains from seven hospitals in China and tested *in vitro* antimicrobial susceptibility. We used Illumina and Nanopore sequencing technology to elucidate the molecular epidemiology and genetic characteristics of the KPC-PA strains.

Our data revealed that the susceptibility to CAZ-AVI of *Pseudomonas aeruginosa* was 50.3% in China. We found that the distribution of resistance genes was mediated by two major KPC-carrying plasmid types. Our data showed that the *bla*_KPC-2_ gene copy number was associated with CAZ-AVI MIC.

## Methods

### Sample collection and antimicrobial susceptibility tests

Clinical CRPA isolates were collected from seven hospitals around China, including from Sir Run Run Shaw Hospital (SRRSH), the First affiliated hospital of Zhejiang University (FAHZU), Provincial People’s Hospital of Zhejiang (ZPPH), Quzhou People’s Hospital (QZPH), Nanjing Drum Tower Hospital (NDTH), Wuhan Tongji Hospital (WTJH) and Peking Union Medical College Hospital (PUMCH). Five hospitals were located in Eastern China (SRRSH, FAHZU, ZPPH, QZPH and NDTH), while other two (WTJH, PUMCH) were in Central and Northern China, respectively. All of these samples were isolated from patients between January, 2017 to February, 2018 and sent to SRRSH for investigation.

The *bla*_KPC_ gene was screened by PCR using the primers KPC-2_FW (5’-AGGTTCCGGTTTTGTCTC-3’) and KPC-2_RV (5’-AGGTTCCGGTTTTGTCTC-3’).

*In vitro* antibiotic susceptibility of 13 antipseudomonal antimicrobial agents was determined by broth microdilution or agar dilution method. The antibiotics included piperacillin-tazobactam, ceftazidime, cefepime, ceftazidime-avibactam, aztreonam, imipenem, meropenem, amikacin, gentamycin, tobramycin, ciprofloxacin, levofloxacin and colistin. Antimicrobial agents were prepared from commercially available sources. Breakpoints were referred to Clinical and Laboratory Standards Institute (CLSI) M100 [20]. *P. aeruginosa* strain ATCC 27853 and *K. pneumoniae* strain ATCC700603 was used as the quality control. Difficult-to-treat resistance (DTR) is defined as *P. aeruginosa* exhibiting non-susceptibility to all of the following: piperacillin-tazobactam, ceftazidime, cefepime, aztreonam, meropenem, imipenem-cilastatin, ciprofloxacin, and levofloxacin [21].

### Whole-genome sequencing

For each KPC-PA strain, a single clone was inoculated into 5 mL Luria-Bertani broth and incubated in a 37□ shaker overnight. Bacteria were lysed by FastPrep-24™ 5G bead beating grinder (MP Biomedicals, CA, USA) at 6.0m/sec for 40 seconds twice. Genome DNA were extracted using QIAamp DNA Mini Kit (Qiagen, Hilden, Germany) according to the manufacturer. DNA concentration was quantified using a NanoDrop 2000 spectrophotometer (Thermo Scientific, Waltham, MA), and verified by agarose gel electrophoresis. Libraries were prepared using the TruePrep DNA Library Prep Kit V2 for Illumina (Vazyme Biotech, Nanjing, China), and sequenced on an Illumina X Ten platform (Illumina Inc., San Diego, CA, USA) and 150-bp paired end reads were generated. The Illumina sequence data were *de novo* assembled by shovill v1.1.0 [22] with options “--mincov 10 --minlen 200 --trim” and SPAdes v.3.13-v.3.14 [23] as the assembler.

To further investigate the molecular characteristics of KPC-PA, we pursued Nanopore MinION long-read sequencing (Oxford Nanopore Technologies, Oxford, UK) on 6 isolates (SRRSH1002, SRRSH1408, NDTH10366, SRRSH15, WTJH12 and QZPH41). These isolates were selected based on their sequence type, plasmid type, *bla*_KPC-2_ copy number and geographic distribution. Another strain, SRRSH1101, *bla*_KPC-2_-negative but harboring Type 1 plasmid, also underwent Nanopore sequencing to help illuminate the dynamics of multiple *bla*_KPC-2_ copies. The raw reads of the sequenced isolates were mapped onto three representative plasmid sequences (GenBank accession: KY296095.1, MN433457.1 and KU578314.1) by bwa-mem v.0.7.17 [24] to identify plasmid type. These plasmids were categorized into three types based on sequence similarity (cutoff is 50%).

### Gene synteny

To attribute contigs to plasmid, genome draft of each strain was aligned to representative plasmid of each type by mummer v.4.0.0beta [25]. Results were filtered with a minimum length of 1000bp. The sum length of each contigs which matched with respective representative plasmid was calculated by a custom R script. Only contigs with more than 50% of length matched with representative plasmid were attributed to plasmid contigs. These plasmid contigs were re-annotated by Prokka v.1.14.6 [26], with representative plasmid as the prior source. Orthologs were found by Orthofinder v.2.4.0 [27]. Gene synteny was visualized by Cytoscape v.3.8.0 [28], according to a published script [29], with minute modification.

### Sequencing depth measurement

To estimate the mean sequencing depth of each strain, SeqKit v.0.13.2 [30] was used to calculate total base amount in reads and genome drafts of each strain. Seqtk v.1.3-r106 [31] was used to subsample short reads randomly to approximately 100× of sequencing depth. Short reads were realigned to genome drafts by bwa-mem v.0.7.17 [24]. To assess the chromosome sequencing depth, single copy gene regions were selected to calculate their average depth with samtools v.1.11 [32] depth with option “-aa”. These single-copy genes were derived from BUSCO v.4.0.0 *Pseudomonadales* ortholog database (10th edition) [33], which including 782 single copy genes. Each strain contained 776 (99.3%) single-copy genes on average. Plasmid sequencing depth were represented by that of *repA* gene. To assess the copy numbers of *bla*_KPC-2_ gene and two types of plasmids, the ratio of sequencing depth of each gene to those of the chromosome were calculated.

### Phylogeny

*P. aeruginosa* genome drafts from NCBI Genbank database were download. The MLST type was detected by mlst v.2.19.0 [34]. Isolates with undefined sequencing type were submitted to PubMLST database [35] for new ST profiles. Orthofinder v.2.4.0 [27] with default options was used to cluster genes into orthologous groups. Sequences of single copy orthologues were aligned by mafft v.7.471 [36] with options (--maxiterate 1000 --localpair). Aligned genes of each strain were concatenated by a custom script to finish the multiple sequence alignment. The best nucleotide substitution models, GTR+I+G4, were selected by ModelTest-NG v.0.1.6 [37] with option (-t ml). The maximum likelihood phylogenetic tree was generated by RAxML-NG v. 1.0.1 [38] with 100 tree searches (50 random and 50 parsimony-based starting trees) and bootstrap replicates (autoMRE criterion). TreeCollapseCL4 [39] was used to collapse branches with <50% bootstrap support into polytomies. A circular tree layout with associated matrix was constructed by the R package *ggtree* v.2.2.4 [40].

### Acquired antimicrobial resistance genes and mutation-derived antibiotic resistance mechanism

Acquired antimicrobial resistance (AMR) genes were annotated by Abricate v.1.0.1 [41], using NCBI AMRFinderPlus database (April 29,2020 updated) [42]. Only antibiotic genes with coverage greater than 90% were included. Pseudomonas-derived cephalosporinase (PDC) and OXA-class β-lactamases were reexamined by BLASTX against amino acid sequences in NCBI AMRFinderPlus database.

To determine mutation-derived antibiotic resistance, a collection of 164 antibiotic-resistance-related genes derived from a previous study was selected as candidates to investigate [43]. Most of previous studies treated model species *P. aeruginosa* PAO1 as the reference. However, natural variations were common among different sequence types. To minimize the false positive results, we chose strains from a previous study as the references for each ST. A published dataset containing both genome sequences and antibiotic susceptibility results was treated as reference [44]. One strain of each ST was selected from meropenem-susceptible group in this dataset and our samples to compose an unduplicated set. According to the accessory genes content analyzed by roary v.3.13.0 [45], each representative strain of each ST in our sample paired with the most similar strain in the meropenem-susceptible group for variant calling. Variants were called by snippy v.4.6.0 [46], and only those in the 164 AMR gene regions were selected. Variants were filtered according to base quality (Phred score) greater than 20 and minimum read depth greater than 20. For *oprD* gene, signal peptide was predicted by SignalP v.5.0 portable version [47]. The *oprD* gene was inferred as nonfunctional if signal peptide is absent or an early stopped mutation occurred.

### Statistical analysis

All statistical analysis were performed using R v.4.0.0-v.4.0.2 and Rstudio v.1.2.5001. Normality were detected by *shapiro*.*test* function in stats package v.4.0.0. Gene copy number comparisons among different MIC groups were done by *dunn*.*test* function in dunn.test package v.1.3.5, between different plasmid type groups were performed by *wilcox*.*test* function in stats package v.4.0.0. Correlation was calculated and tested by *cor, cor*.*test* functions in stats package v.4.0.0, respectively.

## Results

### Geographic Distribution and Antimicrobial susceptibility

A total of 374 CRPA clinical isolates were collected from seven hospitals in China, those were SRRSH (n=86), FAHZU (n=71), WTJH (n=50), PUMCH (n=50), NDTH (n=44), ZPPH (n=35) and QZPH (n=35). 151 strains were *bla*_KPC_-positive based on PCR screens. All *bla*_KPC_ genes were *bla*_KPC-2_. *bla*_KPC-2_ gene were detected in CRPA isolates from all hospitals except PUMCH. The percentage of *bla*_KPC-2_-positive strains in CRPA varied among hospitals from 11.4% (NDTH) to 92.1% (ZPPH) (Figure 1). The antimicrobial susceptibility test on the 151 KPC-PA isolates showed high-level (>90%) resistance to fluoroquinolones and all β-lactams except to CAZ-AVI (Table 1). 93.4% (141/151) of isolates met the criteria of DTR. Half of isolates (76/151) were susceptible to CAZ-AVI. MIC_50/90_ of CAZ-AVI were 16/4 and 32/4 mg/L, respectively. The aminoglycosides resistance rates ranged from 4.0% to 8.6%. No strain was resistant to colistin. Three strains from NDTH were colistin-only-susceptible. Full AST results was shown in Supplementary Table 1.

**Table 1.** *In vitro* Antimicrobial Susceptibility Tests of KPC-producing *P. aeruginosa* from 6 Hospitals in China

| Antibiotics | MIC (mg/L) |  |  | percentage resistance |
| --- | --- | --- | --- | --- |
|  | MIC<sub>50</sub> | MIC<sub>90</sub> | MIC range |  |
| piperacillin/tazobactam | >256 | >256 | 4->256 | 99.34 |
| ceftazidime | 128 | 256 | 4->256 | 100.00 |
| cefepime | >256 | >256 | 2->256 | 99.34 |
| imipenem | >128 | >128 | 8->128 | 100.00 |
| meropenem | >128 | >128 | 1->128 | 100.00 |
| aztreonam | >128 | >128 | 4->128 | 100.00 |
| ceftazidime/avibactam | 16 | 32 | 1->64 | 50.33 |
| amikacin | 4 | 16 | 1->64 | 3.97 |
| gentamycin | 4 | 8 | 0.25->64 | 8.61 |
| tobramycin | 1 | 2 | 0.25->64 | 3.97 |
| ciprofloxacin | >16 | >16 | 0.12->16 | 94.04 |
| levofloxacin | >32 | >32 | 0.25->32 | 93.38 |
| colistin | 0.5 | 0.5 | <0.03-2 | 0.00 |

**Figure 1.**
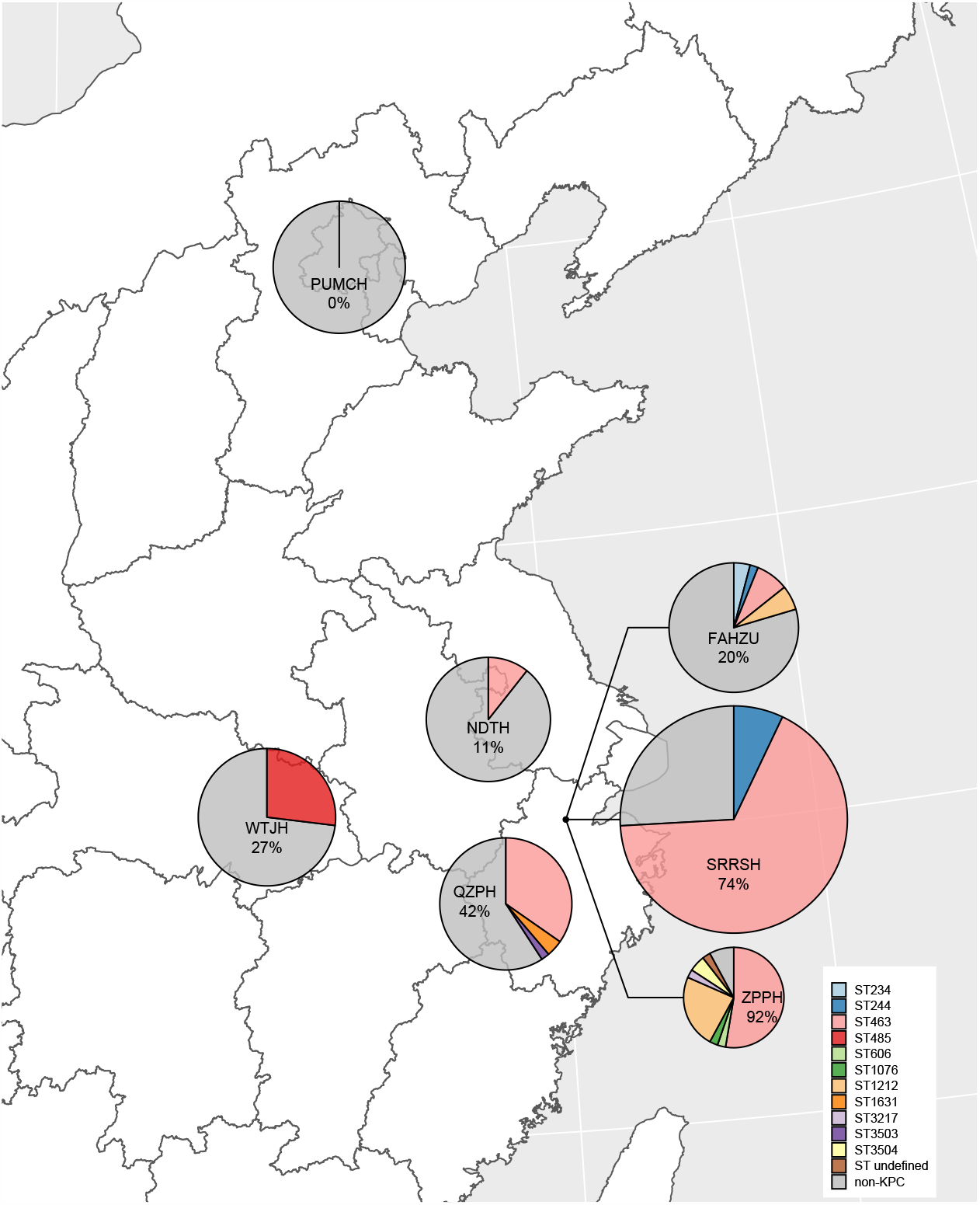
Geographical distribution of strains in this study. The percentages in the figure represent the percentage of KPC-PA in CRPA from each hospital. Each pie size indicates sample size of each hospital. PUMCH: Peking Union Medical College Hospital; WTJH: Wuhan Tongji Hospital; NDTH: Nanjing Drum Tower Hospital; FAHZU: the First Affiliated Hospital of Zhejiang University; SRRSH: Sir Run Run Shaw Hospital; ZPPH: Provincial People’s Hospital of Zhejiang; QZPH: Quzhou People’s Hospital.

### Molecular epidemiology

Making use of Illumina sequencing data available for 151 KPC-PA, we found that the multi-locus sequence type (MLST) detection indicated three main KPC-PA sequence types, ST463 (n=107), ST485 (n=14) and ST1212 (n=12), dominating in different geographic regions (Figure 1). ST463 was mainly found in cities in East China (Nanjing (5/5), Hangzhou (85/113) and Quzhou (17/19)), while ST485 (14/14) is the main KPC-PA of Wuhan in Central China (Figure 1).

### Identification of KPC-encoding plasmids

Making use of sequence alignments, we categorized putative plasmid contigs in each genome draft into three plasmid types. 118 and 19 strains contained Type 1 and 2 plasmids, respectively. One strain, ZPPH8, contained both. No Type 3 plasmid was detected in our samples (Table 2 and Figure 2). The other 15 strains did not contain any of these three plasmid types.

**Table 2.**
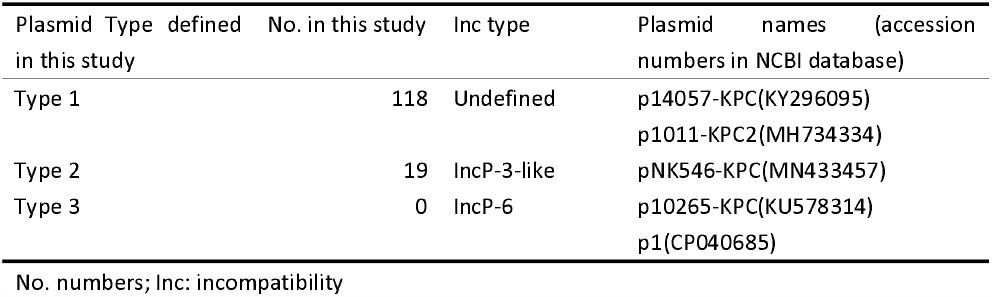
Three main types of KPC-encoding plasmids in *P. aeruginosa* from China

| Plasmid Type defined in this study | No. in this study | Inc type | Plasmid names (accession numbers in NCBI database) |
| --- | --- | --- | --- |
| Type 1 | 118 | Undefined | p14057-KPC(KY296095)<br>p1011-KPC2(MH734334) |
| Type 2 | 19 | IncP-3-like | pNK546-KPC(MN433457) |
| Type 3 | 0 | IncP-6 | p10265-KPC(KU578314)<br>p1(CP040685) |

No. numbers; Inc: incompatibility

**Figure 2.**
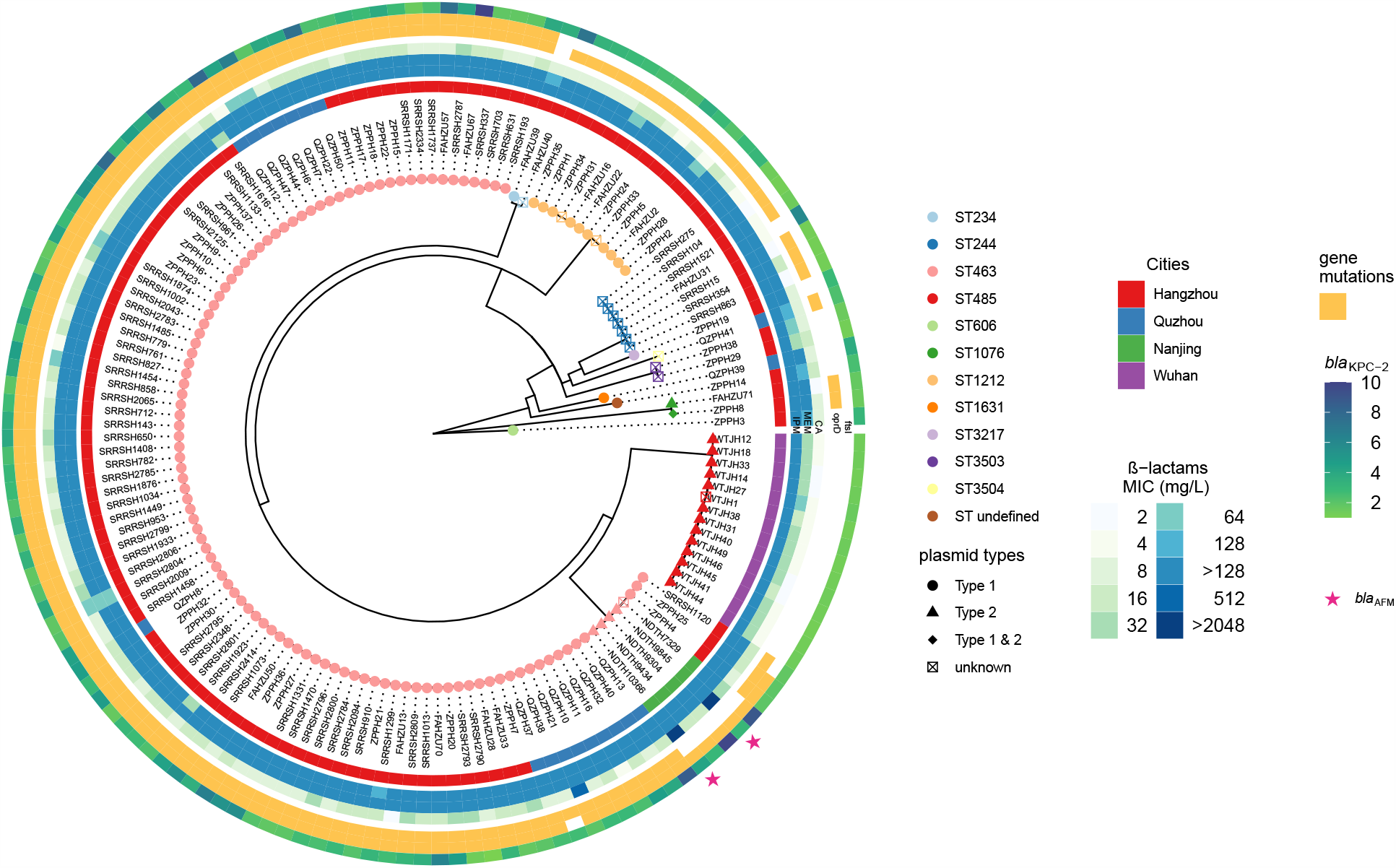
Core genome phylogenetic tree and carbapenem/CAZ-AVI resistance-related genes. Innermost layer is a maximum-likelihood phylogenetic tree of KPC-PA. colors of tip labels indicate sequence type (ST), and shapes indicate plasmid types in each strain. The unknown plasmid type suggests strain does not contain Type 1 or 2 plasmid and *bla*_KPC-2_ gene is located on another replicon. The second ring indicates cities where the strain is isolated. The next three rings indicate MICs of imipenem, meropenem and CAZ-AVI, respectively. The sixth ring represents loss-of-function mutation in oprD gene. The seventh ring represents F533L mutation in PBP3. The eighth ring indicates estimated *bla*_KPC-2_ gene copy number. The stars in the outmost ring indicate novel MBL *bla*_AFM_.

### Molecular characteristics of Type 1 plasmid

A representative of the Type 1 plasmid was pSRRSH1002-KPC. This plasmid was 49,370 bp in length and had a GC content of 58%. Encoded on pSRRSH1002-KPC was a putative repA, a maintenance system and an incomplete type IV secretion system (T4SS) which indicated that this plasmid was not conjugative *per se*. To visualize the structure variation of this plasmid type, we performed a gene synteny analysis which clustered genes of the same orthologs and highlighted the variant part comparing to the reference pSRRSH1002-KPC. From the gene synteny plot, we observed two major variations. One variation we observed was that some members of the Type 1 plasmid kept an intact T4SS. Another group was formed that characterized by gene deletions and inversions in the accessory region (Figure 3A).

**Figure 3.**
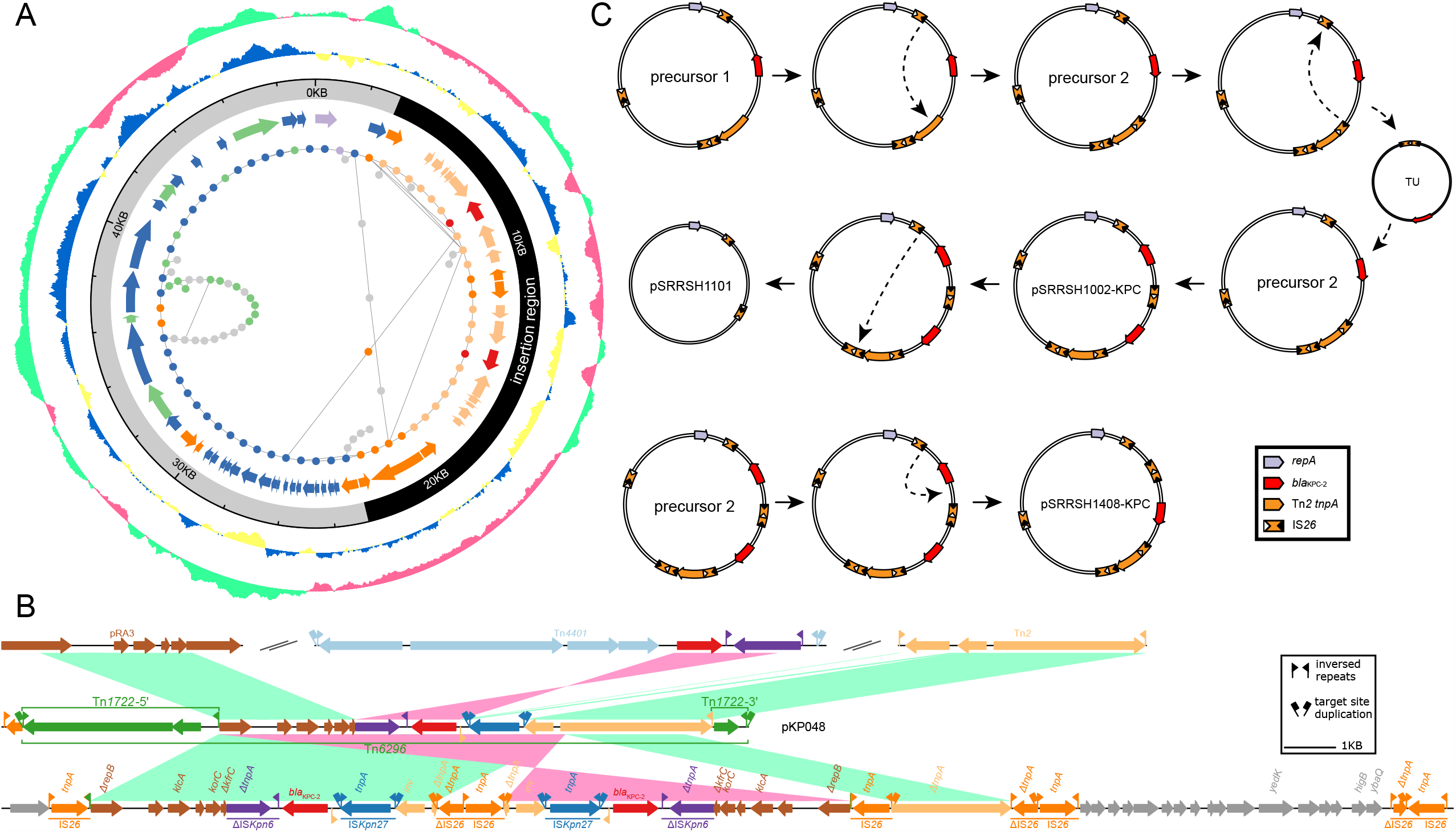
Type 1 plasmid schematic plot and sequence alignment. A. pSRRSH1002-KPC structure and Type 1 plasmid gene synteny. The innermost layer indicates gene synteny of Type 1 plasmids. The arrows in second ring from the innermost indicate genes on SRRSH1002, which are in accordant with points of the gene synteny layers. Orange indicate transposase, red indicates *bla*_KPC-2_ gene, light-orange indicates genes in the insertion region, green indicates conjugal elements, light-purple indicates *repA* gene. The third ring shows backbone(grey) and insertion region(black) of the plasmid. The forth ring represent GC skew, where yellow means skew to A and T and blue means skew to G and C. The outmost ring represents GC content, where light-green represent GC% greater than the median and dark-pink represents GC% less than the median (sliding window of 500bp). **B. Alignment of insertion regions on SRRSH1002-KPC and pKP048.** Arrows denoted to genes or truncated genes. Alignment identity ranges from 97% to 100%. GenBank accession: pRA3 (DQ401103), pKP048 (FJ628167), Tn*4401* from pCOL-1 (KC609323), Tn2 (AY123253). **C. IS_26_-mediated *bla*_KPC-2_ gene copy number variation in Type 1 plasmid**. Firstly, start from Precursor plasmid contained an IS*26*-flanked *bla*_KPC-2_ region. An intramolecular replicative transposition in trans inversed the segment between the 81th bp site on the ΔTn2 tnpA gene and an IS*26* and duplicated an IS*26* in opposite orientation (precursor 2). Intramolecular replicative translocation in *cis* created a translocatable unit (TU). The TU inserted into precursor 2 by conservative targeted transposition to form pSRRSH1002-KPC. pSRRSH1002-KPC excised TU containing the repeated genetic segments and the ΔTn2 *tnpA* gene to form pSRRSH1101. Another plasmid precursor 3 excised a TU containing a *bla*_KPC-2_ gene by intramolecular replicative translocation in cis to form pSRRSH1408-KPC.

Noticeably, pSRRSH1002-KPC harbored two *bla*_KPC-2_ gene copies. The two *bla*_KPC-2_ adjacent regions laid in an inversed repeated pattern, bounded by IS*26*. Each segment was identical to that on pKP048, the most common prototype of *bla*_KPC-2_ adjacent region in China (Figure 3B) [48]. Most often, the flanking insertion sequence was that of IS*26*-ΔTn*6296*. Tn6296 consisted of a core *bla*_KPC-2_ platform, Δ*repB*-*orf1*-*klcA*-*orf2*-*korC*-Δ*kfrC*-ΔIS*Kpn6*-*bla*_KPC-2_-IS*Kpn27*-Tn*2 tnpR*-ΔTn*2 tnpA*, which inserted into Tn*1722* [16]. The Tn*2 tnpA* gene was truncated at the 2,456 bp site. Furthermore, an IS*26* intramolecular replicative transposition in *trans* truncated the Tn*2 tnpA* at the 81 bp site and inverted a 6,922-bp segment as verified by an 8-bp target site duplication (TSD, CGATATTT). Interestingly, different to other IS*26*-franked tandem repeat arrays, both repeated segments were separated by a ΔIS*26* and an intact IS*26*. The first 294 bp of ΔIS*26* was deleted.

We sequenced other two more plasmids, pSRRSH1101 and pSRRSH1408-KPC. pSRRSH1101 was 29,640-bp in size. It had the identical backbone of pSRRSH1002-KPC. However, in pSRRSH1101, the insertion region that contained *bla*_KPC-2_ gene was deleted (Figure 3C). pSRRSH1408-KPC was identical with pSRRSH1002-KPC with the exception of an IS*26* -mediated deletion of one *bla*_KPC-2_ gene (Figure 3C).

### Molecular characteristics of Type 2 plasmid

pNDTH10366-KPC and pWTJH12-KPC belonged to a megaplasmid family[49, 50], referred as Type 2 plasmid here. As with other family members, these two plasmids carried a ParB-related ThiF-related cassette (PRTRC system), a chemotaxis operon, a partition system, a type II secretion system, a radical SAM operon, a tellurite resistance operon and a T4SS on its backbone. The gene synteny plot illustrated the backbone was stable and variable region was composed of different mobile genetic elements containing multiple AMR genes including β -lactamases *bla*_KPC-2_, *bla*_AFM_ and *bla*_OXA-246_ (Figure 4).

**Figure 4.**
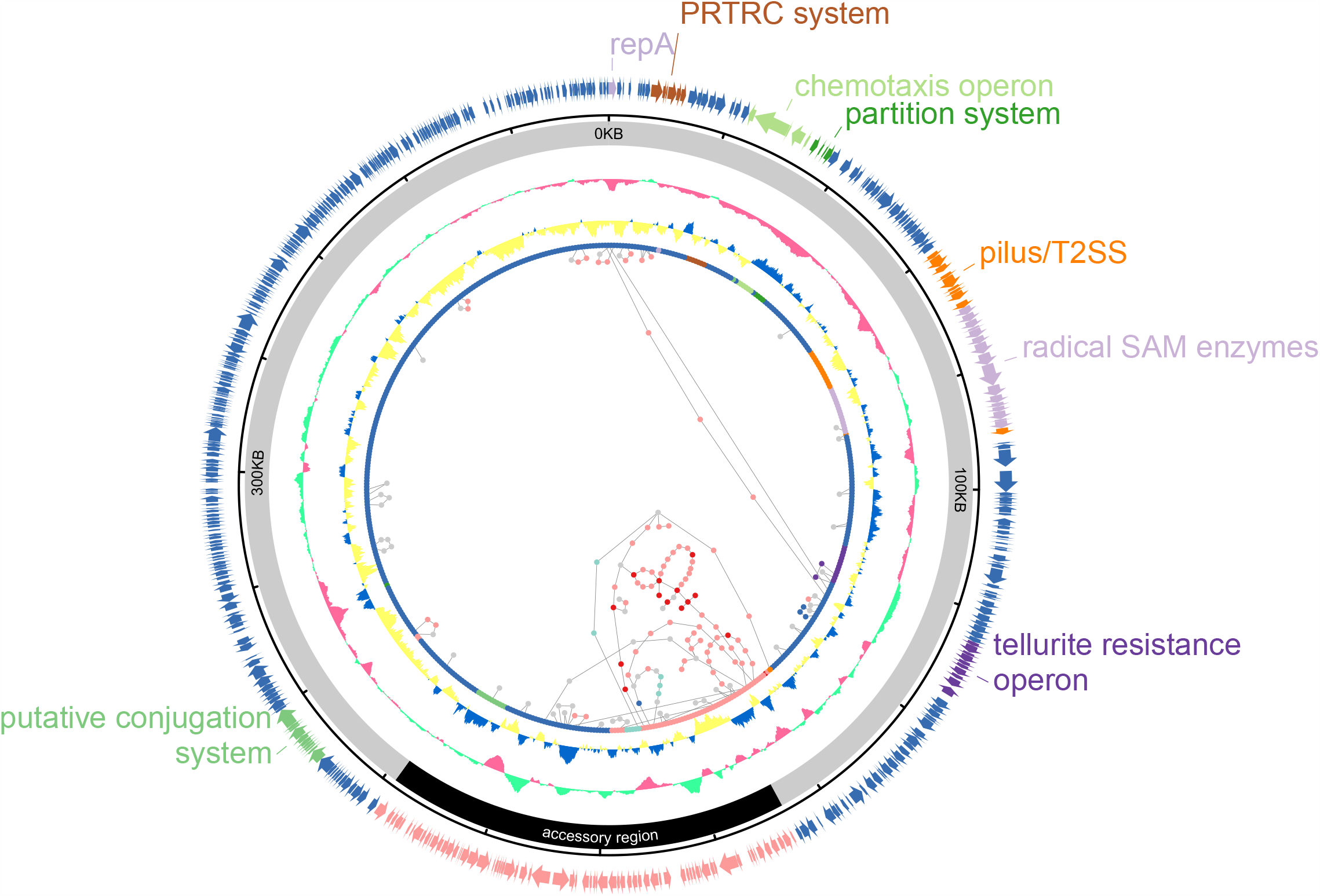
pWTJH12-KPC plasmid structure and gene synteny. The innermost layer indicates gene synteny of Type 2 plasmids. The second ring represent GC skew, where yellow means skew to A and T and blue means skew to G and C. The third ring represents GC content, where light-green represent GC% greater than the median and dark-pink represents GC% less than the median (sliding window of 2000 bp). The forth ring shows backbone (grey) and accessory region (black) of the plasmid. The arrows in outermost ring indicates genes on pWTJH12-KPC, which are in accordant with points of the gene synteny layers. PRTRC: ParB-related ThiF-related cassette; SAM: S-adenosylmethionine; T2SS: Type II secretion system.

### IS*6100*-related composite transposon mediated *bla*_KPC-2_ gene transmission

pWTJH12-KPC contained one *bla*_KPC-2_ gene. The genetic context adjacent to *bla*_KPC-2_ was identical to that on the Type 1 plasmid with p14057-KPC as a representative; in both plasmids the gene is embedded in IS*6100*-ΔTn*6296*-Tn*1403* [16]. This suggested that this mobile genetic element (MGE) carrying *bla*_KPC-2_ might be able to transfer between two plasmid types, especially in case of ZPPH8 which contained both plasmids. The Type 2 plasmid containing the genetic context in which the *bla*_KPC-2_ was embedded, had been recently reported in Tianjin, China [18]. Interestingly, a sequence alignment showed that plasmid pNK546-KPC underwent large segment inversion next to the *bla*_KPC-2_ region (Figure 5). The similarities within the genetic contexts indicated frequent exchange events and transmission of *bla*_KPC-2_-containing genome segments among the strains we investigated, despite of their different sequence types.

**Figure 5.**
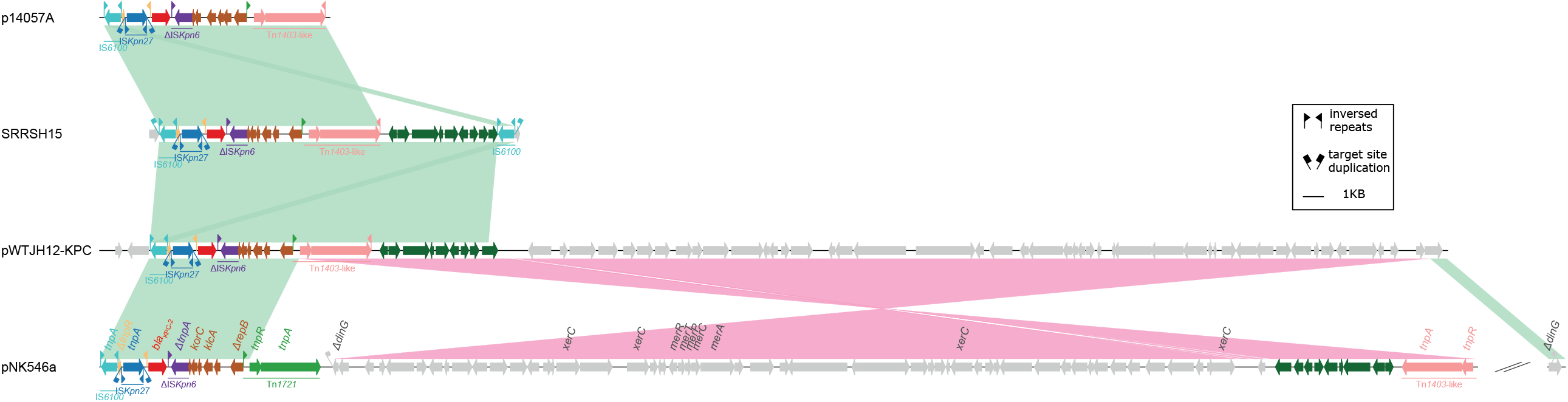
Alignment of IS*6100*-ΔTn*6296*-Tn*1403* structure from multiple strains. Arrows denoted to genes or truncated genes. Alignment identity ranges from 99% to 100%. GenBank accession: p14057-KPC(KY296095), pNK546a (MN433457).

### Comparison of two KPC-encoding plasmid types

Since most of the *bla*_KPC-2_ gene were located on plasmids, we hypothesized that plasmid types and plasmid copy numbers might contribute to the *bla*_KPC-2_ gene multiple copy in each strain. To verify our hypothesis, Illumina sequencing depths of all sequenced isolates were measured. Compared to Type 2 plasmid, Type 1 plasmid exhibited 1.2 more copies per cell (2.36 vs. 1.20, p=1.5×10^−10^, see Table 3). Strains containing Type 1 plasmid harbored 1.4 more *bla*_KPC-2_ gene on average than those containing Type 2 plasmid (2.56 vs. 1.15, p=1.07×10^−7^, see Table 3).

**Table 3.** Comparison between strains containing Type 1 and Type 2 plasmids

|  | Type 1 (n=117) | Type 2 (n=17) | p value |
| --- | --- | --- | --- |
| plasmid copy | 2.36 (1.93-2.98) | 1.2 (1.15-1.29) | 1.45E-10 |
| *bla*<sub>KPC-2</sub> gene copy | 2.56 (2.03-3.49) | 1.15 (1.06-1.30) | 1.07E-07 |

The medians are shown. In brackets indicate interquartile ranges. p values are calculated by Wilcox test.

### *bla*_KPC-2_ gene on chromosome

Although most *bla*_KPC-2_ genes were located on plasmids, we identified two KPC-PA strains harbored *bla*_KPC-2_ gene on the chromosome.

In the *P. aeruginosa* strain NDTH10366, a *bla*_KPC-2_ gene array was found on the chromosome which consisted of three IS*26*-mediated tandem-repeat *bla*_KPC-2_ adjacent segments. Strains from SRRSH that belonged to ST244, also contained a chromosome-embedded *bla*_KPC-2_ gene. The genome of a representative strain SRRSH15 consisted of a single circular 6.7-Mb-long contig. An IS*6100*-flanked composite transposon with a size of 17,797 bp inserted into the chromosome at the site of a putative hydrolase gene, leaving 8-bp TSD (GGCAAGCC) (Figure 5). These findings indicated intracellular transposition occurred in *P. aeruginosa* and participated in shaping *P. aeruginosa* genomes.

### The effect of the *bla*_KPC-2_ copy number and other AMR genes on CAZ-AVI susceptibility

In our samples, all strains exhibited high-level resistance to carbapenems while the susceptibility to CAZ-AVI varied in a large range (2-512mg/L). We further investigated the correlation between CAZ-AVI MIC values and multiple *bla*_KPC-2_ gene copies mediated by both plasmid multiple copies and IS26 or IS6100 duplicative transposition. We excluded two strains containing MBLs (NDTH7329 and NDTH10366) when analyzing the effect of the *bla*_KPC-2_ copy number on CAZ-AVI susceptibility. The *bla*_KPC-2_ gene copy number correlated with the CAZ-AVI MIC values (Pearson coefficient 0.326, 95% CI 0.174-0.463, p<0.0001). Significant differences of the *bla*_KPC-2_ gene copy number were observed among CAZ-AVI MIC groups (2-32mg/L, Figure 6A). However, when values of the high-level resistance(≥64mg/L) were observed, the relationship was less obvious, possibly due to the comparably fewer number of strains exhibiting high-level resistance to CAZ-AVI (n=6).

**Figure 6.**
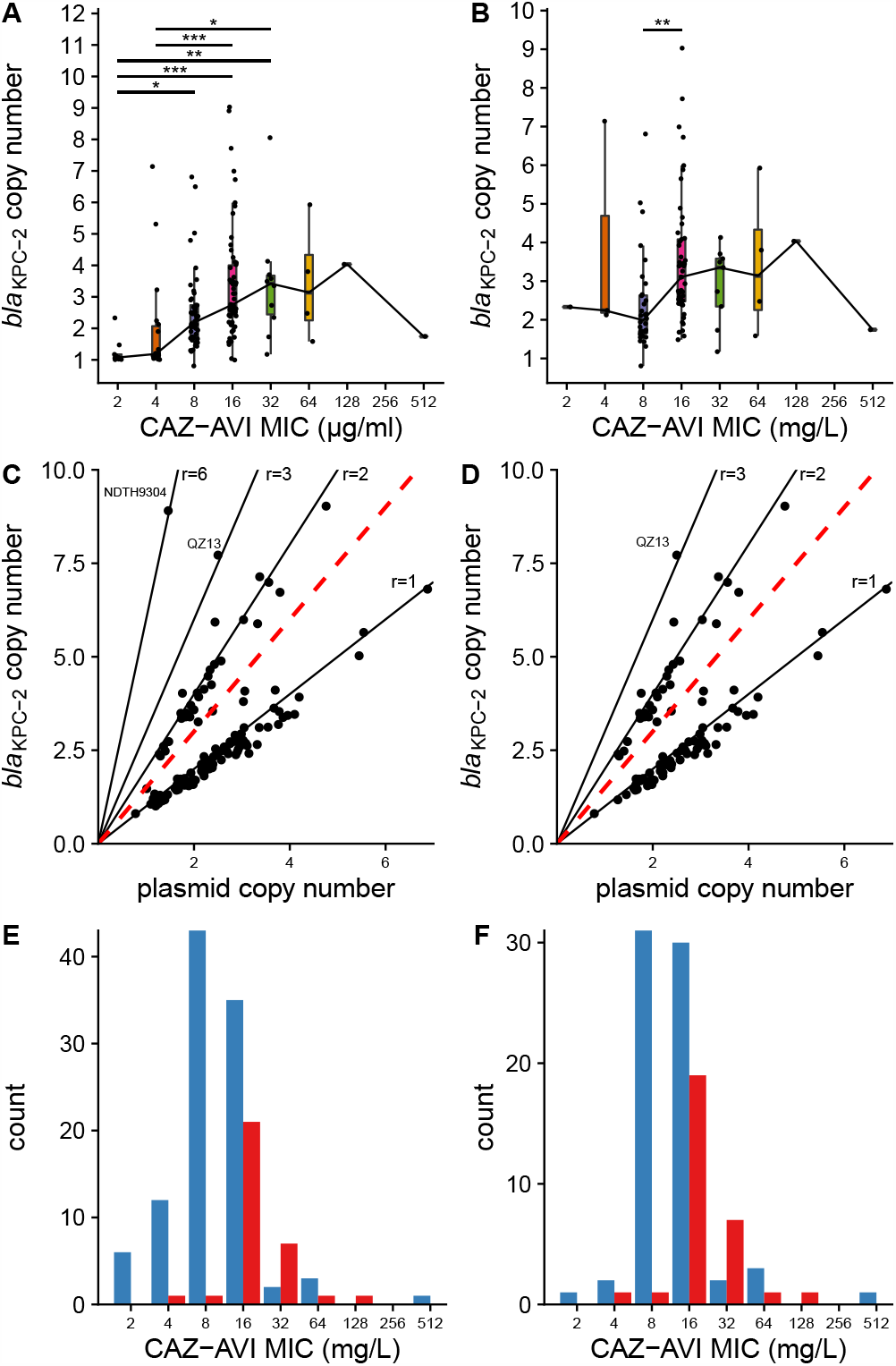
Statistical analysis of the correlation between *bla*_KPC-2_ gene and CAZ-AVI MIC. **A**. *bla*_KPC-2_ gene copy number comparison among strains (n=149) with different CAZ-AVI MIC groups. The asterisks, *, **, ***, represent adjusted p-values of Dunn test less than 0.05, 0.01, 0.001, respectively. Two strains containing metallo-β-lactamase are not involved. **B**. *bla*_KPC-2_ gene copy number comparison among ST463 strains (n=100) with different CAZ-AVI MIC groups. The asterisk, **, represents adjusted p-values of Dunn test less than 0.01. **C**. Correlation between *bla*_KPC-2_ gene copy number and plasmid copy number in strains containing Type 1 or Type 2 plasmid (n=134). **D**. Correlation between *bla*_KPC-2_ gene copy number and plasmid copy number in ST463 strains containing Type 1 plasmid (n=100). Each solid line indicates that points near it exhibit a *bla*_KPC-2_ gene to plasmid copy number ratio as the slope (r). The red dashed line with a slope of 1.5 distinguishes single and multiple *bla*_KPC-2_ gene copies per plasmid. **E**. The CAZ-AVI MIC distributions of strains (n=134) containing single *bla*_KPC-2_ gene or multiple *bla*_KPC-2_ gene copies per plasmid. Both types of plasmids are included. **F**. The CAZ-AVI MIC distributions of strains (n=100) containing single *bla*_KPC-2_ gene or multiple *bla*_KPC-2_ gene copies per Type 1 plasmid. Blue and red bars represent the Single and the Multi group, respectively.

Besides the acquisition of *bla*_KPC-2_ gene, other chromosomal mutations also contributed to carbapenem and CAZ-AVI resistance. The *oprD* gene encoded the outer membrane porin D which allowed the diffusion of carbapenems into *P. aeruginosa*. In this study, all *oprD* genes could be classified into 18 types based on amino acid sequences similarity. 124/151(82.1%) strains were presumed to contain a nonfunctional *oprD* gene, mediating reduced sensitivity to carbapenems. The majority of nonfunctional *oprD* genes were found in ST463, ST244 and ST1212 strains. All *oprD* genes in the clinical isolates from WTJH and NDTH exhibit no mutations, indicating the expression of functional proteins (Figure 2 and Supplementary Table 2).

Carbapenem and CAZ-AVI resistance could also be mediated by the F533L mutation in the *ftsI* gene which encoded penicillin-binding protein 3 (PBP3) (Figure 2 and Supplementary Table 2). In 107 isolates that belonged to ST463 strains, we found almost all of them (n=105) contained a T1597C nucleotide mutation in the *ftsI* gene, which lead to the F533L substitution in PBP3. All twelve ST1212 isolates displayed a so far uncharacterized mutation in the *ftsI* gene (A271G) and correspondingly T91A amino acid substitution in PBP3 which has not yet been studied regarding potential carbapenem or CAZ-AVI resistance mechanisms.

Since these variables such as mutations in PBP3 or porin D might contribute to the observed resistance values besides the acquisition of *bla*_KPC-2_ gene. In order to analyze the contribution of each variable to CAZ-AVI resistance, we analyzed the impact of *bla*_KPC-2_ gene copy number in a more homogenous genetic background. We selected a subset which consisted of 100 ST463 strains with the F533L substitution in PBP3 and nonfunctional porin D, while harboring the Type 1 plasmid. In this subset, significant difference was only detected between 8 and 16mg/L groups (p=0.002, Figure 6B). Again, it might be explained by fewer samples in other groups.

We also investigated the effect of multiple *bla*_KPC-2_ genes copies on a single plasmid to CAZ-AVI susceptibility. We used the ratio of *bla*_KPC-2_ gene sequence depth to the *repA* gene sequence depth as the relative *bla*_KPC-2_ gene copies per plasmid. We defined the ratio less than 1.5 as single *bla*_KPC-2_ gene per plasmid (Single) and that greater than 1.5 as multiple *bla*_KPC-2_ gene copies per plasmid (Multi) (Figure 6C and 6D). In 134 strains containing Type 1 or Type 2 plasmid, the CAZ-AVI MIC values were significantly different between the Single and the Multi groups (p=1.6e-7, see Table 4). This result was still hold in the ST463 subset mentioned above (p=5.8e-5, see Table 5). In both subsets, the plasmid copy numbers were not significantly different. Of note, the Multi group showed a right-shifting tendency of the CAZ-AVI MIC distribution compared to the Single group (Figure 6E and 6F). These results indicated that multiple *bla*_KPC-2_ gene copies caused by insertion-sequence-mediated duplication contributed to CAZ-AVI MIC elevation.

**Table 4.** Comparison between single and multiple *bla*_KPC-2_ gene copies per plasmid groups in 134 strains containing Type 1 or 2 plasmids

|  | Single (n=102) | Multi (n=32) | p value |
| --- | --- | --- | --- |
| log<sub>2</sub>(CAZ-AVI MIC) | 3 (3-4) | 4 (4-5) | 1.63E-07 |
| plasmid copy | 2.32 (1.72-2.90) | 2.07 (1.75-2.45) | 3.60E-01 |

The medians are shown. In brackets indicate interquartile ranges. p values are calculated by Wilcox test.

**Table 5.** Comparison between single and multiple *bla*_KPC-2_ gene copies per plasmid groups in 100 ST463 strains

|  | Single (n=70) | Multi (n=30) | p value |
| --- | --- | --- | --- |
| log<sub>2</sub>(CAZ-AVI MIC) | 4 (3-4) | 4 (3-4) | 5.80E-05 |
| plasmid copy | 2.39 (1.94-3.03) | 2.09 (1.78-2.49) | 0.1003 |

The medians are shown. In brackets indicate interquartile ranges. p values are calculated by Wilcox test.

Other carbapenem or CAZ-AVI resistance related genes including *Pseudomonas*-derived cephalosporinase (PDC) and efflux pumps especially the MexAB-OprM. PDC was a constitutional β-lactamase in *P. aeruginosa* that was able to hydrolyze cephalosporins. PDCs were found to correlate with the sequence types. The *bla*_PDC-8_, *bla*_PDC-6_, *bla*_PDC-3_ genes were identified in three main STs, ST463, ST485 and ST1616, respectively. No amino acid substitutions which might contribute to CAZ-AVI resistance were detected in our samples. No so far known-of-function mutations in efflux pump genes was detected. In addition to *bla*_KPC-2_, acquired β-lactamases observed in individual strains were *bla*_CARB-2_, *bla*_OXA-10_ family β-lactamase *bla*_OXA-246_ and a novel MBL *bla*_AFM_.

Comprehensive AMR-related mutations or acquired AMR genes were listed in Supplementary Table 2.

## Discussion

In this study, we focused on a specific subpopulation of CRPA, that was KPC-PA. At the point of time when the bacteria were isolated, the KPC-PA strains showed moderate susceptibility to CAZ-AVI. The MICs of approximately 70% of the strains were at the margin of breakpoint (8/4-16/4 mg/L). In our samples, we identified two main KPC-related plasmids. Type 1 plasmid was predominant; however, most were found in ST463 strains and their distribution was limited in Eastern China. In contrast to this, Type 2 plasmid was spread more widely and existed in many diverse sequence types. Deploying Nanopore long-read sequencing, we deciphered *bla*_KPC-2_ genetic characteristics in this work. We found that the *bla*_KPC-2_ copy number variation was caused by mobile genetic elements, in particular by IS*26*, mediating transposition. Our results also revealed that multiple copies of the *bla*_KPC-2_ gene correlated with elevated CAZ-AVI MIC values.

Previous epidemiological surveillance studies had focused on CRPA. One of the largest studies performed in China reported a resistance rate of CRPA to CAZ-AVI was 34.3% in 2017 [51] and 35.7% in 2018 [52]. The strains investigated in our study were collected around the same time (2017-2018). The isolated KPC-PA strains exhibited a higher CAZ-AVI resistance rate than all the CRPA. In the previous studies, no screening was performed regarding the *bla*_KPC-2_ gene, which in retrospect showed the unawareness of which important role this AMR gene played in *P. aeruginosa*. As for the origin of *bla*_KPC-2_ gene in *P. aeruginosa*, it was fair to infer that the gene was introduced into *P. aeruginosa* by interspecies transmission since the genomic environment of the *bla*_KPC-2_ gene was near identical and closely related to the gene that has first been described in *K. pneumoniae*. Data from the above-mentioned study [53] indicated that ∼65 % carbapenem-resistant *K. pneumoniae* in China carried the *bla*_KPC-2_ gene, which made it the most prevalent carbapenemase in this species. Interestingly, 100% of *bla*_KPC-2_-carrying *Enterobacteriaceae* were susceptible to CAZ-AVI [53], which stood in contrast to 50% susceptibility in KPC-PA which we observed in this study. It suggested that the acquisition of *bla*_KPC-2_ impacted the CAZ-AVI susceptibility in *P. aeruginosa* in more substantial extent than in *Enterobacteriaceae*. In general, it becomes clear that screening for *bla*_KPC_ in CRPA was important.

Our study also investigated the impact of *bla*_KPC-2_ copy number variation. It was reasonable to assume that there was a correlation between the *bla*_KPC-2_ copy number and CAZ-AVI resistance. Previous studies had demonstrated β-lactamase gene amplification correlated with the susceptibility to β-lactam/β-lactamase inhibitor combination agents [54-56]. Our results supported these conclusions. An important mechanism of AMR gene amplification was insertion-sequence (IS)-mediated duplication. In our study, *bla*_KPC-2_ was located on two mobile genetic elements, IS*26*-ΔTn*6296* and IS*6100*-ΔTn*6296*-Tn*1403*. IS*26*-mediated module appeared to promote genetic element transposition, for example tandem or inversed repeats and from plasmid to chromosome. IS*26* was strongly linked to horizontal AMR gene transfer [57]. Two mechanisms of IS*26* movements had been demonstrated previously, that was the replicative transition and the conserved targeted transition [57, 58]. The characteristic 8-bp TSD can help to trace IS*26* transposition. From our data, we could conclude that the *bla*_KPC-2_ copy number variation among strains harboring Type 1 plasmid was mediated by the conserved targeted transition. In later statistical study, we found that IS-mediated gene amplification contributed to CAZ-AVI MIC elevation. It caveated that the antimicrobial susceptibility test was necessary to surveil during the treatment course, since the MIC might fluctuate due to IS-mediated copy number variation.

Five *P. aeruginosa* plasmids carrying *bla*_KPC-2_ reported in China had been identified on the NCBI database (Table 2). Type 1 plasmid defined in this study was reported first by Shi et al [16] and later by Hu et al [17]. In this work, we provide high-precision sequencing data of three more Type 1 plasmids with different *bla*_KPC-2_ gene copy numbers. Type 2 plasmids belonged to a megaplasmid family which had been reviewed recently [49, 50]. Members of this family displayed a size of 300-500 Kb and several AMR genes. However, those carrying *bla*_KPC-2_ genes were only reported in China [18, 59]. The current study showed that this megaplasmid spread widely around the country as we identified the plasmid in Eastern and Central China. The third plasmid type, reported in other publications, was not detected in this study, indicating that this plasmid type might be not prevalent in regions we sampled. However, two previously reported cases of the third plasmid type were found in China in two different regions of the country [59], which suggested that the third plasmid type was not rare. Fifteen strains in the current study had undefined *bla*_KPC-2_ gene location (chromosome or plasmid). The previously published data and our current work illustrates that a more comprehensive surveillance study was required in the future to elucidate the epidemiology of KPC-PA in China.

The two main plasmid types exhibited different geographical distributions. This might be explained by the time that has passed since the plasmids were incorporated by *P. aeruginosa* strains. Type 1 plasmids had been found in *P. aeruginosa* for approximately one decade [13], while the earliest strain harboring Type 2 plasmid could be traced back to the 1980s [50]. In addition, the host range of Type 1 plasmid was comparatively small, while the Type 2 plasmid family had been found in other Pseudomonal and non-Pseudomonal genera [50]. This might be explained by the fact that the conjugal ability of Type 2 plasmid has been demonstrated experimentally [18], in contrast to Type 1 plasmids. However, the high pressure of carbapenem usage in clinics might accelerate the occurrence of *P. aeruginosa* harboring *bla*_KPC-2_ genes and retain such strains in the nosocomial environment, leading to dominant clones such as ST463.

The limited sampling locations of the strains, which originated mainly from Eastern China, presents a limitation of our study, as the real prevalence of KPC-PA over the whole country remained uncertain. As discussed above, our investigation could be seen as a pilot study and thus we highly recommended to screen for *bla*_KPC_ genes in nationwide antimicrobial surveillance studies, or those being undertaking by other nations. Another potential limitation was that we determined plasmids in each strain by mapping sequencing reads to representative plasmids. We understood that this approach did not guarantee to identify correctly on which replicon the *bla*_KPC-2_ gene was located. However, we believed that it was reasonable to assume that strains of the same ST were highly similar in the genetic background to the representative strains that we sequenced completely. Indeed, this approach had also been implemented in a recent large-scale carbapenemase-harboring plasmid analysis [60].

## Conclusions

In summary, our study clearly shows that KPC-PA strains represent a threat to the healthcare system in China. Therefore, we propose to screen *bla*_KPC_ genes in *P. aeruginosa* isolates in nationwide surveillance projects. As this issue is not only affecting China but is of global concern, further studies investigating the same topic on a global scale would help to understand the epidemiology of KPC-PA strains in the entire human population. We believe such studies might be able to guide therapeutic deploying CAZ-AVI for the treatment of KPC-PA infections.

## Data Availability

All the sequence data were deposited at DDBJ/ENA/GenBank under the BioProject accession number PRJNA672835.

## Declarations

### Ethics approval and consent to participate

Approval was obtained from the Ethics Committee of Sir Run Run Shaw Hospital (approval/reference number: 20201118-49). This study was not considered as a human research. Therefore, no informed consent to participate was required. This study conformed to the principles of the Helsinki Declaration.

### Consent for publication

Not applicable

### Authors’ contributions

Y.Y conceived, designed, and coordinated this study. Y.Z. and J.C. performed the microbiological cultures of the isolates and antimicrobial susceptibility tests. Y.Z and X.H. analyzed the genome sequencing data. H.S., Z.C., Q.Y., J.Z., X.L, Q.Y. and F.Z. collected the isolates from respective hospitals. J.J., H.C., Y.L. and L.Z. provided help to extract genome DNA and perform genome sequencing. Y.Z. wrote the initial version of the manuscript. X.H., S.L. and Y.Y. revised the manuscript. All authors read and approved the final manuscript.

## Acknowledgments

Not applicable

